# *IFIH1* loss-of-function predisposes to inflammatory and SARS-CoV-2-related infectious diseases

**DOI:** 10.1101/2023.10.13.23297034

**Authors:** Rania Najm, Lemis Yavuz, Ruchi Jain, Maha El Naofal, Sathishkumar Ramaswamy, Walid Abuhammour, Tom Loney, Norbert Nowotny, Alawi Alsheikh-Ali, Ahmad Abou Tayoun, Richard K. Kandasamy

## Abstract

The *IFIH1* gene, encoding melanoma differentiation-associated protein 5 (MDA5), is an indispensable innate immune regulator involved in the early detection of viral infections. Previous studies described MDA5 dysregulation linking it to weakened immunological responses, and increased susceptibility to microbial infections and autoimmune disorders. Monoallelic gain-of-function of the *IFIH1* gene has been associated with multisystem disorders, namely Aicardi-Goutieres and Singleton-Merten syndromes, while biallelic loss of this gene causes immunodeficiency. In this study, nine patients suffering from different cases of recurrent infections, inflammatory diseases, severe COVID-19, or multisystem inflammatory syndrome in children (MIS-C) were identified with putative loss-of-function *IFIH1* variants by whole exome sequencing. All patients revealed signs of lymphopenia and an increase in inflammatory markers, including CRP, amyloid A, ferritin, and IL-6. One patient with a pathogenic homozygous variant c.2807+1G>A was the most severe case showing immunodeficiency and glomerulonephritis. The c.1641+1G>C variant was identified in the heterozygous state in patients suffering from periodic fever, COVID-19, or MIS-C, while the c.2016delA variant was identified in two patients with inflammatory bowel disease or MIS-C. Expression analysis showed that PBMCs of one patient with a c.2016delA variant had a significant decrease in *ISG15*, *IFNA* and *IFNG* transcript levels, compared to normal PBMCs, upon stimulation with poly(I:C), suggesting that MDA5 receptor truncation disrupts the immune response. Our findings accentuate the implication of rare monogenic *IFIH1* loss-of-function variants in altering the immune response, and severely predisposing patients to inflammatory and infectious diseases, including SARS-CoV-2 related disorders.

## Introduction

The melanoma differentiation-associated protein 5, MDA5, is a critical element of the innate immune system and is essential for the detection of viral infections. MDA5 is encoded by the *IFIH1* gene and belongs to RIG-I-like receptors (RLRs) that recognizes viral double stranded RNA in the cytoplasm of infected cells. Two N-terminal caspase activation and recruitment domains (CARDs), followed by a helicase, Pincer, and C-terminal domain-like regions (CTD), make up the structure of MDA5. The helicase domain includes RecA-like domains, namely Hel1 and Hel2, and possess ATPase activity needed to unwind double-stranded RNA complexes. The Pincer domain is responsible for the ATPase core activation and structural support, whereas CTD is for dsRNA binding^1–3^. Upon MDA5 activation, the receptor oligomerizes and interact with the mitochondrial anti-viral-signaling protein (MAVS), an adaptor for RLR signal transduction pathway. This is followed by the activation of interferon regulatory factor 3/7 (IRF3/IRF7) and NF-_κ_B transcription factors; hence, inducing the expression of type I and III interferon (IFN) genes^2,4^. Paracrine and autocrine IFN stimulation induce IFN-stimulated genes (ISGs) that are needed for an antimicrobial response ^1,5,6^. This allows the establishment of an antiviral state in the neighbouring cell and regulation of immune cells activation to efficiently mount an antiviral immune response.

Mutations in *IFIH1* have been associated with auto-inflammatory disorders and vulnerability to a wide range of viral infections. Previous studies showed that heterozygous gain-of-function *IFIH1* variants cause a spectrum of auto-inflammatory pathologies including Aicardi-Goutieres syndrome type 7^7,8^, Singleton-Merten syndrome 1^9^, and IgA deficient systemic lupus erythematosus^10^. The immune-pathologies result from the constitutive or aberrant activation of MDA5 with an enhanced sensitivity to self-derived nucleic acid^11^. Consequently, *IFIH1* gain-of-function variants cause an autoinflammatory state of type I interferonopathy via an enhanced or constitutively active downstream MAVS pathway^12^.

Nevertheless, *IFIH1* loss-of-function variants increased susceptibility to viral and frequent severe respiratory tract infections by drastically impairing the immune response^13–15^. Furthermore, autoimmune and immune-mediated diseases such as multiple sclerosis^16^, type 1 diabetes^17^, sclerosing cholangitis^18,19^ and very early onset inflammatory bowel disease ^20^ have been linked to *IFIH1* loss-of-function. The mechanism behind the development of severe disorders by these risk alleles is yet to be elucidated; however, type I interferon dysregulation may highly contribute to susceptibility to infections and in the initiation of autoinflammatory/autoimmune responses^1^.

We have previously shown that *IFIH1* loss-of-function variants are significantly enriched in patients with high susceptibility to infections^21^. Furthermore, transcriptomic analysis of patients with COVID-19 showed that *IFIH1* expression is significantly altered in this group of patients relative to controls^22^. We therefore hypothesized that *IFIH1* regulates common immune-related pathways which, when altered, can increase predisposition to infections such as SARS-CoV-2.

## Materials and Methods

### Study Cohort

Patients presenting with recurrent infections and/or inflammatory disease were found, by clinical genomic testing, to carry loss-of-function variants in the *IFIH1* gene were included in this study (N = 5). In addition, patients meeting criteria for severe COVID-19^23^ or MIS-C^24^ and subsequently determined to have loss-of-function *IFIH1* variants by whole exome sequencing, were also included (N = 4). This study was approved by the Dubai Scientific Research Ethics Committee-Dubai Health Authority (Study numbers: DSREC-07/2023_06, DSREC-SR-03/2023_08, and DSREC-07/2021_05.

### Clinical and Demographic Information

Deidentified patient information was extracted from electronic medical records at Al Jalila Children’s Hospital or Rashid Hospital (Dubai, United Arab Emirates) where those patients were treated and/or recruited for the study. Data was collected on standardized data collection form. Variables included demographic information, clinical manifestation, blood workup results, severity of the disease, course of illness, diagnosis, treatment, and outcome. After reviewing and evaluating all information, common variables and abnormal results among the patients were highlighted as presented below.

### Genomic Sequencing

Whole exome sequencing was performed as previously described^25^. Briefly, after DNA extraction and fragmentation, coding regions were enriched using the Agilent Clinical Research Exome V2 (CREv2) capture probes (Agilent, USA) and subsequently sequenced (2 × 150 bp) using the NovaSeq system (Illumina, USA) to a minimum average depth of 100X.

### Bioinformatic Analysis

Sequencing data were then processed using an in-house, custom-made bioinformatics pipeline to retain high-quality sequencing reads across all coding regions, and prioritize high-quality variants after annotating with allele frequency (using mainly the Genome Aggregation database, the Greater Middle East variome database, and the Middle East Variation (MEV) database)^26^, predicted protein effects, and presence or absence in disease databases, such as ClinVar and the Human Gene Mutation database (HGMD). All variants reported in the *IFIH1* gene were according to the transcript number: NM_022168.3. Loss-of-function enrichment analysis was performed as previously described^21^.

### PBMC Isolation and Treatment

Peripheral blood was obtained from a healthy volunteer and patient #4 with the heterozygous c.2016delA *IFIH1* variant. Peripheral blood mononuclear cells (PBMCs) were isolated from heparinized peripheral blood using Histopaque-1077 (Sigma) by density centrifugation. Normal PBMCs (nPBMC) and patient 4 PBMCs (P4 PBMC) were rested for 24 hours in RPMI-1640 (Gibco) supplemented with 10% Fetal Bovine Serum (FBS) (Sigma) and antibiotics. For poly(I:C) transfection, 5*10^5^ cells were seeded in 500ul of Opti-MEM (Gibco) in a 24-well plate. Cells were transfected with 10µg/ml of poly(I:C) using Lipofectamine 3000 Transfection Kit (Invitrogen) according to the manufacturer’s protocols and stopped after 6, 10, and 24 hours for sample collection. Transfected cells without poly(I:C) and collected after 24 hours were used as negative control. Experiments were performed in 4 biological replicates.

### Gene Expression Studies

Total RNA from normal or patient PBMCs treated or untreated with poly(I:C) was extracted by Trizol (Ambion, Life Technologies). cDNA was synthesized using a Reverse Transcription Kit (Qiagen, QuantiTect). SYBR Green RT-qPCR was carried out using the QuantStudio5 machine. Primer sequences are listed in Table S1. For RT-qPCR, individual reactions included 150ng of cDNA, 0.25 µM of each primer, and PowerUp SYBR Green Master Mix in a final volume of 10_μ_l. PCR reactions started with a DNA denaturation step (95°C, 3 minutes), followed by 40 cycles of denaturation (95°C, 15 seconds), annealing (57°C, 60 seconds), extension (72°C, 30 seconds). Reactions were conducted in duplicate for each experiment. For analysis, individual genes expression was normalized to glyceraldehyde-3-phosphate dehydrogenase (*GAPDH*), a housekeeping gene (Table S1), and Livak method was used to calculate the transcript expression levels^27^.

## Results

### Patient cohort

To further understand the role of *IFIH1* in predisposition to infections, we searched for loss-of-function variants in this gene in three patient cohorts: 1) 51 pediatric patients referred to clinical genomic testing for recurrent infections, periodic fever, immunodeficiency and/or inflammatory disease, 2) 45 patients meeting the criteria for MIS-C, and 3) 55 young (mean age 34 years) patients with severe COVID-19 who were otherwise healthy.

### Identification of *IFIH1* loss-of-function variants

Whole exome sequencing was performed on all 151 patients (62% males) and analysis was restricted to predicted loss-of-function variants in the *IFIH1* gene. Such variants were identified in 9 patients, including two patients diagnosed with severe COVID-19, two patients with MIS-C, three patients with periodic fever, and two other patients presenting with immunodeficiency or inflammatory bowel disease (IBD) (**Table 1**). The same variant, c.1641+1G>C, was identified in 3 patients while the frameshift c.2016delA variant was identified in 2 other patients. Variants were heterozygous in 8 patients, while in patient 6, presenting with variable immunodeficiency and glomerulonephritis, the c.2807+1G>A was in the homozygous state.

**Table 1:** List containing the gender, clinical presentation, *IFIH1* variant and *IFIH1* zygosity of patients included in this study.

| Patient | Gender | Presentation | *IFIH1* variant | Zygosity |
| --- | --- | --- | --- | --- |
| 1 | Male | Periodic fever | c.1641+1G>C; p.? | Heterozygous |
| 2 | Male | Periodic fever | c.1764delA;<br>p.A589Lfs*16 | Heterozygous |
| 3 | Male | Periodic fever | c.769+3A>G; p.? | Heterozygous |
| 4 | Male | MIS-C | c.2016delA;<br>p.D673Ifs*5 | Heterozygous |
| 5 | Male | MIS-C | c.1641+1G>C; p.? | Heterozygous |
| 6 | Male | Immunodeficiency with<br>glomerulonephritis | c.2807+1G>A; p.? | Homozygous |
| 7 | Female | Inflammatory bowel disease<br>(IBD) | c.2016delA;<br>p.D673Ifs*5 | Heterozygous |
| 8 | Male | Severe COVID-19 | c.1641+1G>C; p.? | Heterozygous |
| 9 | Male | Severe COVID-19 | c.2730C>A;<br>p.Cys910* | Heterozygous |

We noticed at least a 5-fold enrichment of loss-of-function variant in the *IFIH1* gene in this combined cohort (N = 151) relative to healthy individuals of Middle Eastern origin (N = 990) (10/302 or 3.3% vs 13/1981 or 0.66%; p = 0.0003, Fisher’s exact test)^28^. This observation confirms previous findings and further implicate a role for *IFIH1* in susceptibility to infections^14,21^.

### Clinical and Laboratory Characteristics of patients with *IFIH1* Loss-of-function

Most patients (median age: 9 years) were male (8/9 or 88.9%) and the majority were Arab descendants (8 Arabs and 1 Bangladeshi) (**Table 2**). Despite the small size of this *IFIH1* cohort, the sex distribution suggests a possible association between *IFIH1* variants and susceptibility to infections in males.

**Table 2:** Summary for the patients’ demographic information (gender, background, and age) and blood workup results (absolute lymphocyte, CRP, ESR, Amyloid A, Ferritin, and IL-6). CRP: C-Reactive Protein, ESR: Erythrocyte Sedimentation Rate.

|  |  |
| --- | --- |
| **Gender** | 8/9 males |
| **Ethnicity** | 8/9 Arabs |
| **Age (yrs)**<br>**Median (range)** | 9 (1.5-30) |
| **Absolute Lymphocyte (4.00-9.00) 10(3)/μL**<br>**Median (range), N** | 1.2 (0.5-5.6), 9 |
| **CRP (0-2.8 mg/dl)**<br>**Median (range), N** | 123 (13-324), 8 |
| **ESR (<10)**<br>**Median (range), N** | 53 (23-100), 5 |
| **Amyloid A (n<6.4 mg/L)**<br>**Median (range), N** | 475 (263-486), 3 |
| **Ferritin (6-67 ng/ml)**<br>**Median (range), N** | 393 (144-1750), 5 |
| **IL-6 (n< 7 pg/ml)**<br>**Median (range), N** | 109 (32-1362), 3 |

All 9 patients in this cohort presented with fever and signs of autoimmune or inflammatory disease. The initial clinical picture on presentation for each patient was suggestive of viral or bacterial infections, then the course of illness indicated immune system malfunction. Blood workup across this cohort revealed high inflammatory markers including C-reactive protein (CRP), erythrocyte sedimentation rate (ESR), ferritin, and interleukin 6 (IL-6) (**Table 2**).

Increased Amyloid A in patients 1, 2, and 3 indicated a chronic inflammation that could be secondary to autoimmune or chronic infection^29^ (**Table 2**). Glomerulonephritis in patient 6 and Inflammatory Bowel Disease in patient 7, both have immune-related or autoimmune backgrounds^30,31^. Dysregulated immune response to SARS-CoV-2 in previously healthy patients led to hyperinflammation and cytokine storm in patients with severe COVID-19 (patients 8 and 9) or MIS-C (patients 4 and 5)^32^. All patients received immunosuppressive medication such as corticosteroids.

### Expression analysis

The role of homozygous c.2016delA;p.D673Ifs*5 MDA5 variant has been previously described in IBD^20^. In our study, the heterozygous form is detected in two patients 4 and 7 diagnosed with MIS-C and IBD, respectively. The putative truncated MDA5 receptor resulting from c.2016delA variant may lack essential domains including, Hel2, Pincer, and CTD (**Fig. 1A**). Thus, to decipher the role of MDA5 receptor in the severity of infection with SARS-CoV-2, cytokine and interferon gene transcript levels were evaluated in the PBMCs derived from patient #4. The screened cytokines included *IFNA, IFNB, IFNL1, IFNG, IL-6, IL-10 and IL-1B* in addition to the interferon gene transcript *ISG15*, where only transcripts with significant variation are shown.

**Figure 1:**
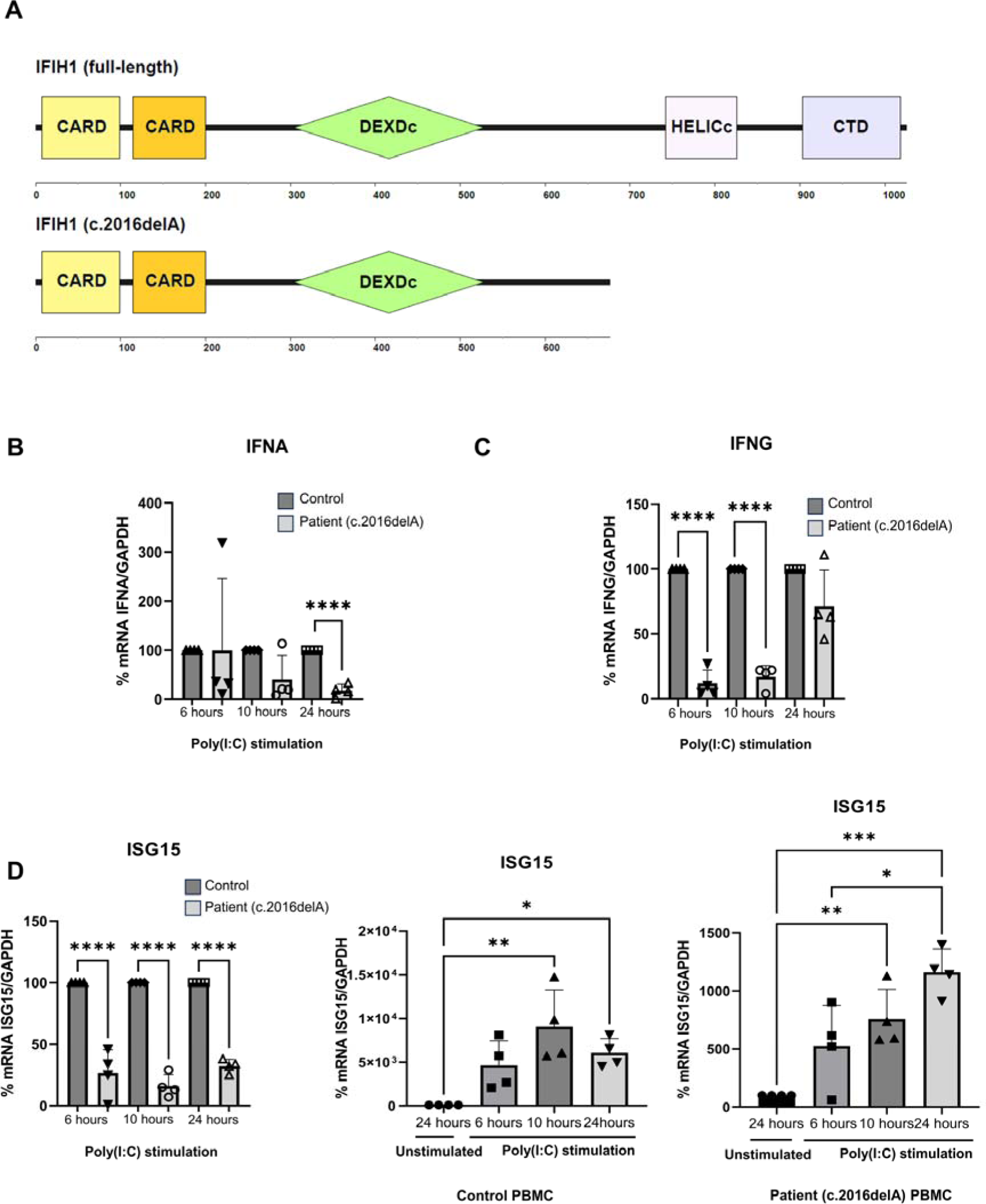
Decreased levels of *IFNA, IFHG*, and *ISG15* transcripts in peripheral blood mononuclear cells with p.D673Ifs*5 MDA5 truncation. (A) A scheme presenting the domains of the full length MDA5 protein versus the potential truncated from of IFIH1 c.del2016A. CARDS: Caspase Activation and Recruitment Domains, DEXDc: DEAD and DEAH box helicase domain, HELICc: Helicase C Domain, CTD: C-terminal domain-like regions. Normal PBMCs (nPBMC) and patient 4 PBMCs (P4 PBMC) were transfected poly(I:C) for 6, 10, and 24 hours. Transfected cells without poly(I:C) were collected after 24 hours. RT-qPCR for *IFNA*, *IFNG* and *ISG15* expression was performed and normalized to *GAPDH*. The results are expressed as percentage of treated nPBMC (±) SD for (A) *IFNA*, (B) *IFNG* and (C-left panel) *ISG15*. The results are expressed as percentage of untreated nPBMC (±) SD for (C-middle panel) *ISG15* and untreated P4 PBMC for (C-right panel) *ISG15*. The student’s t-test was performed to validate significance. *, **, and *** indicate p-values ≤ 0.05; 0.01; and 0.001, respectively.

We used poly(I:C) to simulate RNA virus infection and studied its effect on truncated MDA5 activation in mononuclear cells. Since MDA5 is a cytoplasmic receptor, PBMCs isolated from a healthy individual and patient #4 were transfected with poly(I:C) for 6, 10 and 24 hours, to ensure delivery to the cytoplasm, or without poly(I:C) for 24 hours. Interestingly, *IFNA* transcript level significantly decreased by 82% at 24 hours (**Fig. 1B**) and type II *IFNG* at 6 and 10 hours by 88% and 83% (**Fig. 1C**), respectively, post stimulation in patient PBMCs comparted to normal cells. Moreover, *ISG15* expression was significantly reduced at 6, 10 and 24 hours by 73%, 84%, and 68% (**Fig. 1D**), respectively, post transfection with poly(I:C) in patient comparted to normal PBMCs. Nonetheless, *ISG15* transcript level was still significantly increased in stimulated patient PBMCs at 6, 10 and 24 hours compared to non-stimulated condition at 24 hours (**Fig. 1D**).

These results suggest that the function of the putatively truncated form of MDA5 appears to be disrupted based on the decrease in the expression of type I *IFNA*, and type II *IFHG* and interferon-stimulated gene *ISG15*.

## Discussion

Autoinflammation, chronic inflammation, autoimmunity, and high susceptibility to microbial infections are signs of immune dysregulation, caused by environmental triggers, genetic predisposition, or infections^33^. Mutations in the pathogen recognition receptors, which are responsible for detection of microbial components, alter the innate immune response, hence inducing either an inadequate or self-targeted immunity. MDA5 is an essential innate RIG-1-like receptor that mounts a type 1 innate immune response against viral infections. Genetic alterations of MDA5 leading to loss-or gain-of-function are translated to a wide range of immune diseases. In our study, we highlighted the consequences of *IFIH1* putative loss-of-function variants in altering the immune response and predisposing patients to viral infections like SARS-CoV-2, inflammation, and immune-related disorders.

In this study, we showed that six different *IFIH1* variants were detected in nine patients suffering from severe COVID-19, MIS-C, periodic fever, inflammatory bowel disease, or immunodeficiency with glomerulonephritis. Interestingly, different patients with varied *IFIH1* heterozygous variants had the same clinical manifestation, while the same variant showed different clinical outcomes in different patients. This illustrates the complex phenotypic and genotypic heterogeneity in *IFIH1* related disorders, though environmental factors as well as the genetic background of the patients may also have contributed to the difference in clinical symptoms.

One of the disorders in our cohort is glomerulonephritis, which is the main cause of end-stage and chronic kidney diseases. Susceptibility loci in innate immune genes like *IRF4* and *NFKB1* were related to membrane nephropathy^34^. Herein, one patient had a homozygous *IFIH1* variant, c.2807+1G>A, leading to immunodeficiency with glomerulonephritis. This case was the most severe compared to other cases with heterozygous variants of *IFIH1*, emphasizing the dosage effect on the severity of symptoms and predisposition to diseases.

Periodic fever syndromes are a spectrum of disorders caused by pathogenic variants of the inflammasome and other innate immune related proteins^35^. These include Familial Mediterranean fever gene (*MEFV*)^36,37^, NLR family pyrin domain containing 3 (*NLRP3*)^38^, tumor necrosis factor receptor superfamily 1A (*TNFRSF1A*)^39^, and others. Herein, patients # 1, 2, and 3 with periodic fever were associated with *IFIH1* heterozygous variants. The observation that patients had low lymphocyte count, lymphopenia, and increased Amyloid A suggests that the above variants contribute to chronic inflammation due to chronic infection or autoimmunity hence leading to periodic fever.

MDA5 is known to be essential in inducing antiviral type I and III interferons against SARS-CoV-2 virus^40^. Thus, the severity of the coronavirus disease, COVID-19, is inversely correlated with type I interferon produced upon MDA5 activation^41^. In our study, we reported two cases with severe COVID-19 and the *IFIH1* putative loss-of-function variants c.1641+1G>C and c.2730C>A, which are likely to impair MDA5 activation, hence disrupting type I interferon activity and reducing viral clearance leading to severe COVID-19 outcomes.

In a recent study, a patient identified with very early onset inflammatory bowel disease was characterized with a homozygous variant in *IFIH1* (c.2016delA). The resulting truncated MDA5 (p.D673Ifs*5) receptor, lack Hel2, Pincer, and CTD domains. As a result, the patient suffered from multiple viral and bacterial infection leading to sepsis^20^. Herein, we identify this variant in the heterozygous state in two patients (patients #4 and #7) who presented with different clinical consequences, multisystem inflammatory syndrome in children (MIS-C) and inflammatory bowel disease, respectively^24^. IBD is a consequence of altered immunity towards certain bacterial species and viruses in the gastrointestinal tract which causes inflammation^42^. Patients with homozygous *IFIH1* variant were found to be subjected to bacterial superinfections^13,14^. MDA5 has been proposed to play a role in the activation of immunological responses during bacterial septicaemia^43^. Indeed, Li *et al*. illustrated that RLR/MAVS pathway is activated by commensal bacterial RNA and hence modulates the mucosal immunity towards various microbes in the intestine^44^. As such, MDA5 dysregulation can be linked to inflammatory bowel disease.

In our study the *IFIH1* c.2016delA;p.D673Ifs*5 variant, associated with IBD in this (patient #7) and previous studies, was intriguingly present in patient #4 who was diagnosed with MIS-C. Our expression analysis showed that type I interferon (*IFNA*), type II interferon (*IFNG*), and interferon stimulated genes (*ISG15*) transcript levels were significantly reduced in PBMCs of this patient compared to wild type upon stimulation with poly(I:C) at different time points. Cananzi *et al.* showed that the homozygous p.D673Ifs*5 variant impairs MDA5 ability to activate the *IFNB1* promoter, implying a complete loss-of-function that might result in type I interferon reduction^20^. In addition, deletion or silencing of RIG-1-like receptors (MDA5, LGP2) in iPSC-derived airway epithelium or non-small-cell lung cancer cells led to a decrease in type I IFN expression upon SARS-CoV-2 infection^45,46^.

Studies have highlighted that *ISG15* expression, an ubiquitin-like protein (Ubl), can be induced by type I IFN (IFN-α/β) in a dependent^47,48^ and independent manner prompted by other immune activators^49,50^. Heat-killed Salmonella and poly(I:C) were shown to induce *ISG15* expression in a type I IFN-dependent manner^48^, which explains the reduction in *ISG15* expression in our study. In the non-canonical pathway, extracellular ISG15 binds LFA-1 (Lymphocyte function-associated antigen 1) on surface of lymphocytes, especially NK cell, thereby inducing IFN-γ secretion^48,51,52^. Binding LFA-1 augments IFN-γ as well as other pro-inflammatory cytokines secretion including, IL-1, IL-6, CXCL1 and CXCL5, in addition to an anti-inflammatory cytokine, IL10^51,53^. Hence, the reduction in *IFNG* expression can be linked to the decrease in *ISG15* due to *IFIH1* loss-of-function. On the contrary, infection with SARS-CoV-2 does not only implicate MDA5, but is associated with multiple inflammatory pathways activation causing hyperinflammation and a cytokine storm^54^. Indeed, patient #4 suffered from hyperinflammation which suggests another source of pro-inflammatory cytokine secretion rather than activated MDA5 receptor. In addition, the patient had lymphopenia which might be related to IFNG levels as IFN-γ was shown to mediate lymphocyte and monocyte recruitment to the site of infection^55,56^. Overall, MDA5 dysregulation might lead to the disruption of innate and adaptive immune responses against infections.

In an antiviral defence mechanism, ISG15 induces the ISGylation of certain host and pathogenic proteins inhibiting several viral infections including but not limited to influenza A, HIV, Zika, hepatitis B and C^57^. ISGylation of the viral sensor MDA5 caspase CARDs at Lys23 and Lys43 allow its oligomerization, thus leading to its activation^58^. This activation initiates the antiviral innate immune response that act against a wide range of viruses such as coronaviruses, picornaviruses, and flaviviruses^58^. Conversely, viruses deubiquitinate central signaling proteins like TBK1, IkBα, IKK, RIG-I, and MAVS, via viral deubiquitinating enzymes (DUBs) that might target ISGylation as well to combat immunity^59–61^. Coronaviruses are capable of supressing IFN-mediated antiviral responses, where low IFN levels in SARS-CoV-2-infected patients were associated with severe disease^62^. Moreover, MDA5 ISG15-activiation was shown to be antagonized via de-ISGylation by a papain-like protease (PLpro) of SARS-CoV-2^58^. Herein, our results show a significant increase in ISG15 transcript level in poly (I:C) stimulated PBMCs carrying the putative p.D673Ifs*5 variant compared to non-stimulated cells, which might be explained by a possible translation of MDA5 wild-type form due to heterozygosity. Consequently, aiding in partial but not complete loss-of-function upon stimulation with poly(I:C). The increase in ISG15 in MDA5 mutant PBMCs can also suggest another source of immune activators^49,50^. Nonetheless, this may not imply a partial activation of MDA5 in patient #4. As SARS-CoV-2 PLpro would de-ISGylate MDA5^58^ leading to total functional impairment during infection. This might elucidate the severity of the MIS-C patients case possessing MDA5 mutation, especially that MDA5 is the main SARS-CoV-2 sensor in the lungs^46^.

Additionally, children were observed to have higher expression of RIG-I, MDA5 and LGP2^63^ which can be translated to a pre-activated innate immune response leading to an early onset of interferon production in children’s infected airways^63,64^. This allows a faster viral clearance and lower viral load compared to adults, which might explain the severity of SARS-CoV-2 infection in the MIS-C patient possessing a potential truncated form of MDA5, as these patients are vulnerable to viral infections.

## Conclusion

Pathogenic loss-of-function *IFIH1* variants predispose patients to recurrent infections, autoinflammatory, and autoimmune disorders. Identifying MDA5 pathogenic variants and deciphering the mechanisms and downstream pathways for the initiation of MDA5-related disorder is essential for the development of targeted therapies and interventions to support the immune system in affected patients.

## Data Availability

All data produced in the present study are available upon reasonable request to the authors

## Notes

### Competing Interest Statement

The authors have declared no competing interest.

### Funding Statement

This study was fully funded by grant AJF202059 from the Al Jalila Foundation, Dubai, United Arab Emirates

### Author Declarations

Ethics committee/IRB of Dubai Health Authority gave ethical approval for this work.

